# Healthcare-associated Urinary tract infection and its determinants among Adult Patients Admitted to Intensive Care Units of Addis Ababa Public Governmental Hospitals, Ethiopia; 2020

**DOI:** 10.1101/2023.12.05.23299476

**Authors:** Wondimagegn Genaneh, Tigist Nega, Hindu Argeta, Silenat Gashaw, Eyouel Shimeles

## Abstract

**Back ground:** Urinary tract infections are common bacterial infections that affect almost 150 million people internationally each year. A catheter-associated urinary tract infection arises when germs enter the urinary tract via a urinary catheter, leading to infection and have been linked with increased mortality, morbidity, healthcare costs in intensive care units. It is one of the highest prevalent health care-related infections, accounting for nearly 30% of intensive care unit (ICU) reports because of its association to urinary catheterization, but has great preventive potential.

**Method:** Institutional based cross-sectional study design applied to determine the prevalence and associated factors of Health care-associated urinary tract infections among adult 391 patients admitted to ICU from 2017 to 2019 GC at Addis Ababa Public Governmental Hospital, Addis Ababa, Ethiopia, June-December 2020.Data had manually checked and entered to Epi-data manager version 4.6 and statistical analyses have been performed using SPSS version 23 software program. Strength of association between dependent and independent variables is assessed using crude odds ratio (COR) and adjusted odds ratio (AOR) with confidence Interval (CI) of 95%. Variables that had a value of P < 0.25 on bi-variate analysis were directly forward to be analyzed by multi variable analysis. Goodness of fit test had also computed for logistic regression using Hosmer and Lemeshow test resulted in (sig=0.073), finally having P-values < 0.05 is considered as statistically significant.

**Result:** the study find that the prevalence of Healthcare Associated Urinary Tract Infection among ICU admitted patients was 91(23.3%) 95%CI ;(19.2-27.4), While length of stay, Having tracheostomy, patients on Mechanical Ventilation and taking Proton pump inhibitor drugs were associated with HAUTI in the study area.

**Conclusion:** Healthcare-associated Urinary Tract infection is highly emerging clinical condition among ICU admitted patients in the study areas.

## 1. INTRODUCTION

A catheter-associated urinary tract infection arises when germs enter the urinary tract via a urinary catheter, leading to infection and have been linked with increased mortality, morbidity, healthcare costs in intensive care units [1]. It affects any part of the urinary system, including the kidney, ureter, bladder, and urethra [2].

Although HAIs constitute a major danger in all healthcare settings, their incidence is higher in patients in intensive care units (ICUs) than in patients on other wards [3]. In developed countries, 5–15% of hospitalized patients and more than 50% of patients in intensive care units (ICUs) develop healthcare-associated infections (HCAIs) In resource-limited countries, the magnitude of HCAIs is underrated or unknown due to the absence of well-established surveillance system [4]. Patients admitted to the ICU are predisposed to HAIs owing to reduced body defense, severe underlying disease, poor nutritional status, multiple exposures to invasive devices, and extensive range spectrum of drugs [5].

Urinary tract infections (UTIs) are common bacterial infections that affect almost 150 million people internationally each year. Urinary tract infections (UTIs) are caused by a wide-ranging pathogens, comprising Gram-negative and Gram-positive bacteria, as well as fungi, the most common causal agent for both uncomplicated and complicated UTIs is uropathogenic Escherichia coli (UPEC). [6].

Urinary tract infection (UTI) is one of the highest prevalent health care-related infections, accounting for nearly 30% of intensive care unit (ICU) reports because of its association to urinary catheterization, but has great preventive potential [7, 8]. Nosocomial UTIs have been associated with a threefold increased risk for mortality in hospital-based studies, with estimates of more than 50,000 excess deaths occurring per year in the USA as a result of these infections [9].

During hospitalization, indwelling urethral catheters account for about 80% of urinary of UTIs. Catheters may facilitate colonization of the urinary bladder due to poor catheter placement, prolonged catheterization, poor aseptic technique, poor hand hygiene, and poor asepsis of the urethral orifice opening [10]. Indeed, as reported by ECDC, the urinary catheter utilization rate was 78 per 100 patient-days in ICUs, and nearly 98% of UTIs were associated with the presence of a urinary catheter [6]. Commom risk factors for ICU patients includes Prolonged catheterization, Female gender, Catheter insertion outside operating theatre, patients with sepsis, Urology service, immune-compromised patients, advanced age, prolonged ICU stay, Other active sites of infection, Diabetes, Malnutrition, Creatinine >2 mg/Dl, Ureteric stents, Rigorous monitoring of urine output, Improper positioning of drainage tube and Antimicrobial drug therapy [2] [9, 11]

Serious sequelae such as frequent recurrences, pyelonephritis, colitis cystitis, renal damage, Gram-negative bacteremia, preterm birth endocarditis, vertebral osteomyelitis, septic arthritis, and complications caused by frequent antimicrobial use, such as antibiotic resistance and Clostridium [12]and meningitis in all patients can occur [13, 14]. these lead to discomfort for the patient, with an excess mortality rate of 23 deaths per 1000 inpatients and excess costs of $1000/case, i.e., additional costs per hospital acquired infection [14].

The US Centers for Disease Control and Prevention (CDC) strongly recommends using soapy water, and distilled water or povidone-iodine, for sterilizing the periurethral area before catheter performance; However, the results of previous studies have been contradictory. Some studies have reported that this approach was not effective in decreasing the UTI rate [8]. Shortening the duration of catheterization, avoiding unnecessary catheterization, preparing and implementing standard infections control protocols and using sterile precautions in the insertion and maintenance of indwelling urinary catheters can reduce catheter-related complications [2, 12]

## 2. METHODS AND MATERIALS

### 2.1. Study design, Objective, Population, Area and P eriod

Institutional based retrospective cross-sectional study design was conducted to determine the prevalence and associated factors of Healthcare–associated Urinary tract infection among adult patients admitted from 2017 to 2019 to intensive care unit of Addis Ababa Public Governmental Hospitals June to September 2020

### 2.2. Inclusion and Exclusion Criteria

Adult Patients’ charts admitted Addis Ababa Public Governmental Hospital ICUs from 2017 to 2019 and stayed above 48 hours of hospital admission were included while incomplete or vague charts and having prior diagnosed infection were excluded from the study.

### 2.3. Sample size determination

To calculate the sample size to determine the prevalence and associated factors of HAUTI in the two hospitals were done by single proportion formula:

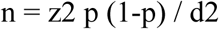

Where; n= sample size

d= maximum allowable error (margin of error) = 0.05

Z = value of standard normal distribution (Z-statistic) at 95% confidence level (z=1.96).

P = Expected prevalence (Proportion) since in our country there is no specific ICU study regarding prevalence and associated factors of Health care-associated Urinary tract infections, p-value of 0.5 considered

n= (1.96)2×0.5 (1-0.5)/ (0.05)2=384 then 5% of sample size is considered for incomplete charts and un-available (contingency), 384(0.05) results 19.2

Total sample size will be 384+19.2= 404

### 2.4. Sampling techniques

Saint Paul’s Hospital Millennium Medical College and its affiliated Addis Ababa Burn, Emergency and Trauma hospital has been select ed by lottery method among all federal hospitals find in Addis Ababa. After proportional allocation of 187 and 217 individual charts to Saint Paul’s Hospital Millennium Medical College and its affiliated Addis Ababa Burn, Emergency and Trauma hospital, systematic random sampling technique was employed to select ICU cases admitted for treatment, to be included as the study subject, which mean, after arranging patients charts sequentially the first patient chart taken as case one by lottery method from the first three patient medical charts. Then the other study subjects were selected and included in every three clients chart intervals.

### 2.5. Operational and term definitions

**Health Care Associated Urinary Tract infection;** is defined as either a microbiologically confirmed symptomatic UTI or non-microbiologically confirmed symptomatic UTI which was developed after 48 hours of ICU admission.

**Catheter-associated urinary tract infection**; is a HAI developed after insertion of urinary catheter, central line catheters and endotracheal or tracheostomy tube all the way to the body either in intermittent or continuous ways within the 48-hour period before onset of infection.

**Multi-drug resistance (MDR)**; an isolated microbe that is not susceptible to at least one antimicrobial agent in three or more antimicrobial classes

### 2.6. Data collection technique

Data were collected from April-01, 2020 to April-22, 2020 by two trained health professionals from patients’ medical records using slightly modified check list adopted from previous studies [31-34]. Simultaneously the principal investigator monitor, clarify and evaluate the entire data collection procedures by following expected ethical procedure. Case definitions of HAIs were adapted from European Center for Disease prevention and Control reporting format (ECDC) [34]

### 2.7. Data quality management

Data collectors had been by two trained health professionals, in which the principal investigator has monitored, clarified and supervised whole activities of the study .Then the collected data checked for the completeness and consistency and then loaded to statistical software in care full manner for both data cleaning and analysis.

### 2.8. Data processing, analysis, presentation and interpretation

After collection the data checked for completeness and coded and entered to Epidata manager version 4.6 by principal investigator. Then, it was exported to SPSS Statistics version 23 for analysis and cleaning.

Descriptive statistics expressed by frequency, percentages and median with Inter quartile range. The associated factors were determined using bivariate and multivariate binary logistic regressions. Normality test has been done using Kolmogorov-Smirnov and Shapiro-Wilk test. Multicollinarity test (Variable inflation factor =1.00) also done to examine similarities between the independent variables. Goodness of fit test had also computed for logistic regression using Hosmer and Lemeshow test resulted in (sig=0.073).

Strength of association between dependent and independent variables is assessed using crude odds ratio (COR) and adjusted odds ratio (AOR) with confidence Interval (CI) of 95%. Variables that had a value of P < 0.25 on bi-variatee analysis were directly forward to be analyzed by multi variable analysis then having P-values < 0.05 is considered as statistically significant. The result is presented by using tables, chart and texts.

### 2.9. Ethical consideration

Ethical clearance was obtained from SPHMMC after informed by written letter, then Permission also been taken from both Hospitals administrations to conduct the study after submitting Original ethical approval letter. Then data have been collected from patient charts in a way that made it impossible at least very hard to identify using newly assigned code numbers and by keeping questionnaires in safe place.

## 3. RESULT

### 3.1. Socio-demographic status of the participants in Addis Ababa Public Governmental Hospitals, Addis Ababa Ethiopia, 2020

From the total sample size Three hundred ninety one (97 %) patients’ were enclosed in the study and the rest participants’ charts were unavailable during the collection. The age of the patients was range between 18 and 94 years and merely half (50.1%) of the participants’ age were grouped between 18-34 years. Thirty four years were the median age of the participants with an inter-quartile range of plus or minus 30 years.

Nearly half of the research participants Two hundred nine(53.5%) were male with male to female ratio of 1: 0.87. Nine days were the median ICU length of stays with an inter-quartile range of 15 days. Patients had been admitted in the ICU between 3 and 104 days. Emergency outpatient was the most referring Hospital department 199 (50.9%) followed by medical ward 70 (17.9%). Only Eighty four (21.5%) participants had previous admission history.

**Table 1.** Socio-demographic status of the participants in the two years period Addis Ababa, Ethiopia.

| Variable | Category of variable | HAUTI |  | Total (%)<br>n=391 |
| --- | --- | --- | --- | --- |
|  |  | No (76.7%)<br>n=300 | Yes (23.3%)<br>n=91 |  |
| Sex | Male | 157(52.3) | 52(57.1) | 209(53.5) |
|  | Female | 143(47.6) | 39(42.8) | 182(46.5) |
| Age group of participants | 18-34 | 147(49) | 49(53.8) | 196(50.1) |
|  | 35-55 | 83(27.6) | 18(19.8) | 101(25.9) |
|  | ≥56 | 70(23.3) | 24(26.4) | 94(24) |
| Length of stay in ICU | 3-9 days |  |  |  |
|  | ≥9 days | 167(55.7) | 17(18.7) | 184(47.1) |
|  |  | 133(44.3) | 74(81.3) | 207(53) |
| Department before to ICU | Emergency OPD | 155(51.7) | 44(48.2) | 199(50.8) |
|  | Medical ward | 53(17.7) | 17(18.6) | 70(17.9) |
|  | Surgical ward | 29(0.1) | 16(17.0) | 45(11.5) |
|  | Obstetrics and gynecology | 41(13.5) | 6(0.1) | 47(12) |
|  | Others sites | 22(0.1) | 8(0.1) | 30(7.6) |
| Previous admission history | Yes | 68(22.7) | 16(17.6) | 84(21.5) |
|  | No | 232(77.2) | 75(82.3) | 307(78.5) |

### 3.2. Clinical characteristics of the study participants at Addis Ababa Public Governmental Hospitals; Addis Ababa, Ethiopia

Respiratory failure 171(43.7%), Traumatic Brain Injury 86(22%), and Acute Kidney Injury 86(22%) were found the commonest underlying disease conditions in the study areas.

About 236 of participant’s baseline Glasgow Coma Scale (GCS) was above eight. About (77.7%) participant had received prophylaxis antibiotics and 279 (71.4 %) study population were intubated while only Thirty six (9.2%) had chest tubes .Merely 12 (1.3%) participants had Central vascular catheters. More than two third 327 (83.6%) of admitted patients had indwelling urinary catheters in situ.

**Figure 1.**
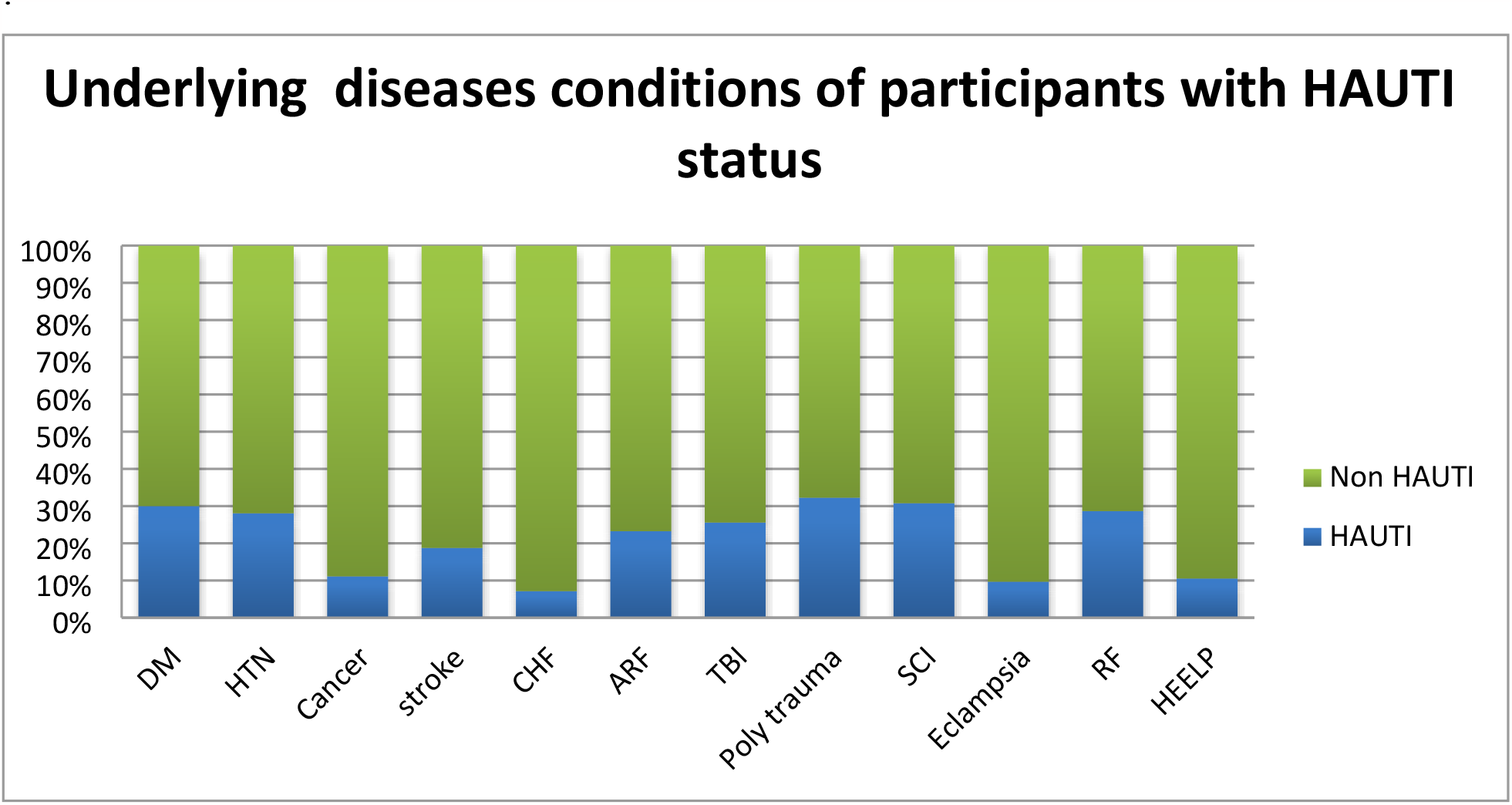
Common underlying diseases with their HAUTI status among adult ICU participants in Addis Ababa Public Governmental Hospitals, Addis Ababa Ethiopia., 2020

**Table 2.** Clinical characteristics of the study participant at Addis Ababa Public Governmental Hospitals, Addis Ababa Ethiopia., 2020.

| Variable | Category of variable | HAUTI |  | Total (391)<br>N(%) |
| --- | --- | --- | --- | --- |
|  |  | Yes(91)<br>n (%) | No(300)<br>n(%) |  |
| **GCS** | 3-8 | 44(48.4) | 111(37) | 155(39.7) |
|  | 9-15 | 47(51.6) | 189(63) | 236(60.3) |
| **Central vascular catheter** | Yes | 21(23.0) | 51(17) | 72(18.4) |
|  | No | 70(77) | 249(83) | 319(81.6) |
| **Endotracheal tube** | Yes | 85(93.4) | 194(64.6) | 279(71.3) |
|  | No | 6(6.6) | 106(35.4) | 112(28.6) |
| **Urinary catheter** | Yes | 87(95.6) | 240(8) | 327(83.6) |
|  | No | 4(4.4) | 60(92) | 64(16.4) |
| **Mechanical ventilator** | Yes | 85(93.4) | 195(65) | 280(71.6) |
|  | No | 6(6.6) | 105(35) | 111(28.4) |
| **Tracheostomy** | Yes | 38(41.7) | 24(8) | 62(15.9) |
|  | No | 53(58.3) | 276(92) | 329(84.1) |
| **Nasogastric tube** | Yes | 84(92.3) | 208(69.3) | 292(74.6) |
|  | No | 7(7.7) | 92(30.7) | 99(25.4) |
| **Chest tube** | Yes | 13(14.2) | 23(7.6) | 36(9.2) |
|  | No | 78(86.8) | 277(92.4) | 355(90.8) |
| **Surgery within a month** | Yes | 50(54.9) | 135(45) | 185(47.3) |
|  | No | 41(46.1) | 165(55) | 206(52.7) |
| **Received prophylaxis Antibiotics** | Yes | 84(92.3) | 220(73.3) | 304(77.8) |
|  | No | 7(7.7) | 80(27.7) | 87(33.2) |
| **Blood transfusion** | Yes | 30(32.9) | 118(39.3) | 148(37.9) |
|  | No | 61(68.1) | 182(60.7) | 243(62.1) |
| **Immune-suppressant** | Yes | 43(47.2) | 117(39) | 160(41) |
|  | No | 48(52.8) | 183(61) | 231(59) |
| **Sedative Drugs** | Yes | 57(62.6) | 117(39) | 174(44.5) |
|  | No | 34(38.4) | 183(61) | 217(55.5) |
| **Proton pump inhibitors Drugs** | Yes | 52(57.1)) | 148(49.3) | 200(51.1) |
|  | No | 39(42.9) | 152(50.7) | 191(48.1) |

Where DM-Diabetes mellitus, HTN-Hypertension, CHF-Congestive Heart Failure, ARF-Acute Renal Failure, TBI-Traumatic Brain Injury, SCI-Spinal Cord Injury and RF-Respiratory Failure, HELLP-Hemolysis Elevated Liver enzyme Low Platelets

### 3.3 Prevalence of health care associated Urinary tract infections with categories at Addis Ababa Public Governmental Hospitals

Total of Ninety one (23.3) 95%CI ;(19.2-27.4) study participants had been diagnosed to have healthcare associated urinary tract infection in the study areas. Approximately 87(95.60 %) of with health care associated Urinary tract infections were associated with Urinary Catheter (CAUTI).

### 3.4 Identified pathogens of HAUTI in the two years period at Addis Ababa Public Governmental, Addis Ababa Hospitals Ethiopia

Almost for 14.8 % participant’s culture test had been performed and fifty bacteria (microorganism) have been identified. Of recognized microbe about 86% were gram negatives bacteria’s. Acinto bacter spp (37%) were the highest number of identified microbe, Subsequent to it klebsiella spp (16%), E-coli (11%), Citrobacter and Proteus spp (11%) were found to highly prevalent causative agents of HAIs. About 72% of isolated bacteria were resistant for more than one antimicrobial

**Figure 2.**
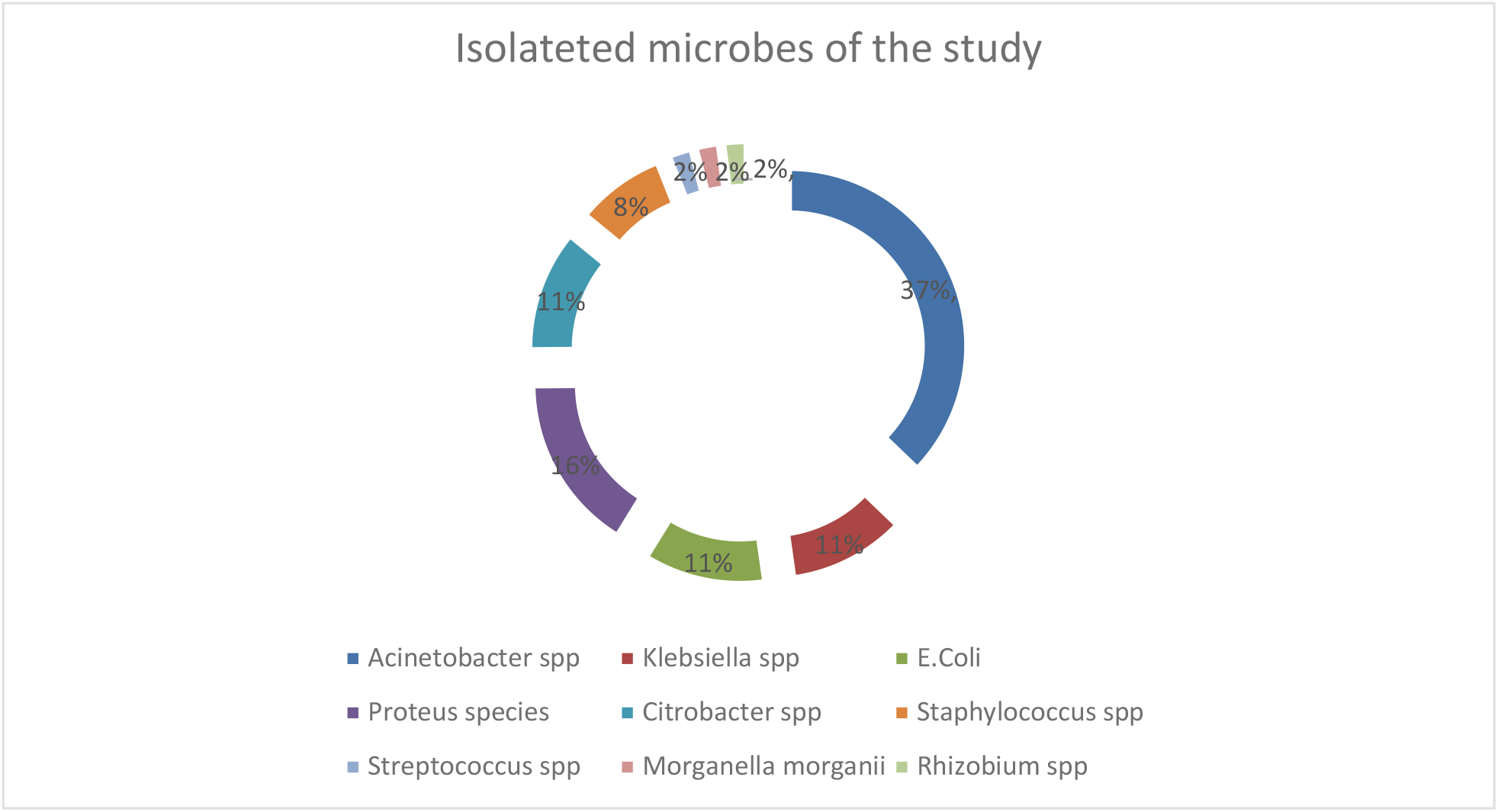
HAUTI distribution within Isolated microorganisms from varies specimens among adult ICU patients at Addis Ababa Public Government Hospital, Ethiopia, 2020 (n=50)

### 3.5 Determinants of Healthcare associated Urinary Tract infections among adult ICU patients at Addis Ababa Public Governmental Hospitals, Addis Ababa Ethiopia

On Bivariate logistic regression analysis the following variables had found to be statistically significant with HAUTIs these variables were; Age, Length of stay, prior to ICU admission, cancer, CHF, Polytrauma, Eclampsia, Respiratory failure, GCS,Central vascular catheter, Endotracheal intubation, Indwelling urinary catheter, Tracheostomy, Chest tube, Nasogastric tube, Surgery within a month, received prophylaxis antibiotics, taking immunosuppressant drug, taking sedative agents, Proton pump inhibitor and Blood transfusion.

However in multivariate binary logistic regression about four variables were obtained to have significant association with HAUTI these were; Length of stay, patients with tracheostomy, patients who took Proton pump inhibitors and patients who was on Mechanical ventilation.

As the Patients length of stay rises by a day the possibility of acquiring infection was increased by 1.032 (AOR=1.032, 95%CI: (1.016, 1.049), P<0.001).Participants with tracheostomy were five times higher risk for HAUTI (AOR=5.33; 95%CI: (2.29, 12.41) P<0.001).

Patients who were with Mechanical ventilation were six time more expected to acquire UTI (AOR=5.61, 95%CI: (1.06-29.65) P=0.043) and taking PPI were 2 times higher possibility of getting HAUTIs (AOR=2.04; 95%CI: (1.03, 4.5), P=0.041).

**Table 3.** Determinants of Healthcare associated Urinary tract infections among patients admitted in Addis Ababa Public Governmental Hospitals, Addis Ababa Ethiopia,, 2020.

| Characteristics | HAUTI<br>Yes | No | COR (95% CI) | P-value | AOR (95%CI) | P-value |
| --- | --- | --- | --- | --- | --- | --- |
| Length of stay<br>in ICU (days ) | 91 | 300 | 1.05(1.03-1.06) | 0.001 | 1.03(1.02,1.05) | 0.001 |
| Tracheostomy |  |  |  |  |  |  |
| Yes | 11 | 21 | 8.25(4.57-14.86) | 0.001 | 5.33(2.28, 12.41) | 0.001 |
| No | 188 | 171 | 1 |  | 1 |  |
| Mechanical<br>Ventilation |  |  |  |  |  |  |
| Yes | 85 | 195 | 7.63(3.22-18.05) | 0.001 | 5.61(1.06-29.65) | 0.043 |
| No | 6 | 105 | 1 |  | 1 |  |
| Proton pump<br>inhibitors |  |  |  |  |  |  |
| Yes | 52 | 148 | 1.37(0.85-2.19) | 0.193 | 2.04(1.028-4.05) | 0.041 |
| No | 39 | 152 |  |  |  |  |

## 4. DISCUSSION

An overall prevalence of Hospital acquired Urinary Tract infection in the two Hospital ICUs was 91 (23.3%) 95% CI ;(19.4-27.6%) and about 95.6% were associated with urinary catheterization under possibly assessed journals the study finding is the first prevalence report in our country which is particular to ICU. This prevalence of HAUTIs in the study area is much higher than findings of Mohd Saleem et al 6.4% [15], a study done in Italy 2.7% [3], 6.5% of Canadian survey [9], Oumer et al.16.8% [14]

The higher prevalence result might be due to consequence inadequate trained health professionals, and inadequate adherence of infection control measures[16]. And also the nature of study participant in ICU; may have reduced body defense, sever comorbid diseases, poor nutritional status, a number of times exposure to invasive devices, extensive range of spectrum of drugs and it is also stated that trauma patients tend to present with higher prevalence[17-19] Whereas the number is closely similar with Aleksa Despotovic MD et al finding with a prevalence of 20.83% [20].

This could be due to ICU service and setting difference, dissimilar management and follow up approaches, severity of comorbid illness and participants’ diverse nature rate.

Gram negative bacteria (86%) specifically Acinto bacter spp (37%), klebsiella spp (16%), E-coli (11%) and Proteus spp (11%) were the main infectious agent in the study which in line with South, Ethiopian Study [14], Saudi Arabia [15], Ugandan [21] Mohammed Saber’s[1],, Indian[22-24], Fiji[25] and Romanian[26] ICU studies. The most dominant bacteria species in this study; Acinetobacter is commonly originated from water supplies of hospitals and contaminated materials [21].

From isolated microbe the rate of Multi-drug resistant (78%) bacteria was very high as compared to Nigerian study (57.1%) and Ugandan study (55.8%) [21, 27] and this may attributed by a very poor ways of devise insertion and maintaining. And is approaches to the finding of study done in Southern Ethiopia (88.1%), it may be due to similarity in region both found.

As the Patients length of stay increases by a day, the likelihood of acquiring infection were increased by 1.03(AOR=1.03, 95%CI: (1.02, 1.05), P<0.001) .It is supported by WHOs HAIs report[28], Yisiak Oumer et al [14], Linchuan Wang and his siblings report[29]. The similarities as length of stay increases, there would be an increment of exposure to hospital environment, exposure to numerous invasive lines including urinary catheter and worsening of comorbidities that increases susceptibility of acquiring infection.

Patients with Tracheostomy were five times higher risk for developing HAUTIs (AOR=5.33; 95%CI: (2.28, 12.41) P<0.001) which is in lined with study done in Thailand [30], The finding may be explained as, patients with Tracheostomy tends to have stay longer in ICU, which may increases exposure to an infection, the tube may restricts their sensory and motor skills and proceed to lessen range of motion as well as by suppressing their gag and cough reflex might disrupt them from safeguarding the airway which may cause aspiration and infection risk may increase from surgical procedure.

Participants with Mechanical Ventilation were 5.61 times higher risk of acquiring HAIs (AOR=5.61; 95%CI: (1.06-29.65), P=.043). Almost similar with the finding of Le-Wen Shao et al and Peter Agaba et al [31, 32]. Mechanical ventilation either through Endotracheal or tracheostomy tube increases the risk of infection from improper procedural application and predisposes hematogenous spread of infection to different areas including Urinary tract.

Patients who took Proton pump inhibitors were 3 times more likely to acquire infection in the hospitals (AOR=2.04, 95%CI: (1.028-4.05), P=0.041). This may be PPI mechanistically, lessening of gastric acidity may lead to amplified gastric passage of pathogens or viable exogenous drug-resistant strains, delayed gastric emptying, increased bacterial translocation, and dysbiosis, resulting in intestinal colonization or infection [33]. This condition may increase the risk of infection especially HAP, which leads UTI through hematogenous spread.

## 5. CONCLUSION

It is established that the prevalence of HAUTI is established to be high in the study areas. Length of stay, Having Tracheostomy, Mechanical ventilation use, taking Proton Pump and Inhibitors were determinants of Health Care Associated Urinary Tract Infections in the study areas.

## 6. RECOMMENDATIONS

### For Federal Ministry of Ethiopia

Healthcare-associated infections rate is one of the most vital indicator of quality of care, it is better to reinforce routine national surveillance of HAIs specially HAUTIs and further monitor implementation practices of alleviating HAUTIs simultaneously encouraged to emphasis on Infection prevention and control practices.

### For Addis Ababa Public Governmental Hospitals

It is suggested to evaluate evidence based practices regarding Mechanical ventilation, drug administration, Tracheotomy care furthermore by facilitating investigation and management process it is better to shorten patient’s length of stay in hospitals, specially in ICUs.

Patients having tracheotomy and on Mechanical Ventilation may requirements strict aseptic technique, evidence based practice and further daily monitor subsequently afterward.

### For health professionals

Since the rate of HAUTIs may show quality of comprehensive care, it is necessary for the intensive care Nurses, Physician and other professionals working on patient care to build up their role on Infection Prevention practices.

### For Researchers

It is crucial to conduct further studies to identify other key determine of HAUTIs might be better to conduct prospective study. And also it is recommended to observe cause-effect relationship of some outlandish findings like higher likely hood of acquiring Urinary tract infection among Proton Pump Inhibitor.

## Data Availability

All relevant data are within the manuscript and its Supporting Information files.

## ACRONYMS AND ABBREVIATIONS

CA-UTI: Catheter-Associated Urinary Tract Infections
CVC: Central Venous Catheter
ECDC: European Centre for Disease prevention and Control
HAIs: Health Care-Associated Infections
HAUTI: Health Care-Associated Urinary Tract Infections
HAP: Hospital Acquired Pneumonia
ICU: Intensive Care Unit
IPC: Infection Prevention Control
MDR: Multi Drug Resistant
Nis: Nosocomial Infections
RICU: Respiratory Intensive Care Unit
SPHMMC: St. Paul’s Hospital Millennium Medical College
UTI: Urinary Tract Infection

